# Hippocampal transcriptome-wide association analysis of shared genetic risks between posttraumatic stress disorder and Alzheimer’s disease

**DOI:** 10.1101/2025.08.11.25333445

**Authors:** Eric Du, Jing Tian

**Affiliations:** Blue Valley West High School, Overland Park, Kansas; Department of Pharmacology and Toxicology, University of Kansas, Lawrence, Kansas

**Author notes:** Corresponding authors, Corresponding authors: Jing Tian, PhD, Postdoctoral Researcher, Department of Pharmacology and Toxicology, The University of Kansas, Lawrence, KS 66045.

## Abstract

The etiopathogenesis of Alzheimer’s disease (AD) is puzzled by the heterogeneous nature of this neurodegenerative disorder. In recent years, clinical and basic research has accentuated a relationship between post-traumatic stress disorder (PTSD) and AD risk. Despite several pathways related to neuroinflammation, metabolic disturbances, and stress, emerging evidence implicates genetic risks at the nexus of PTSD and AD. However, the genetic link between the two conditions is relatively understudied. In this study, we adopted tissue-specific transcriptome-wide association studies (TWAS) with a special emphasis on the hippocampus, a shared vulnerable brain region between AD and PTSD to investigate common genetic risks between the two brain disorders. By leveraging large-scale GWAS summary statistics from large-scale AD and PTSD cohorts, we applied FUSION TWAS and identified susceptibility genes common to both disorders. Further functional annotation mounted TWAS-identified cross-disease susceptibility genes to multiple pivotal biological pathways, especially those related to cell metabolism. Metabolic pathway analysis topped lipid metabolism-related pathways such as “acyl-CoA hydrolysis” that overlaps AD and PTSD risk. The simple interpretation of our results is that AD and PTSD share common susceptibility genes. Deregulation of these common susceptibility genes in the hippocampus may potentiate PTSD and AD in sequence through hippocampal dysmetabolism. These findings from computational analysis shed light on the genetic association between AD and PTSD, which will endorse further investigation through experimental approaches for a better understanding of the etiopathogenesis of AD and PTSD as well as the link between the two conditions from a perspective of precision medicine.

## Introduction

Characterized by insidious onset and progressive cognitive impairment, Alzheimer’s disease (AD) constitutes one of the leading causes of death in the elderly[1]. However, the etiopathogenesis of this devastating neurodegenerative disorder so far remains elusive. In addition to the well-documented interactions between aging process and toxic protein aggregates in AD pathogenesis[1, 2], a large body of evidence has accentuated other risk factors and health conditions that predispose older adults to the development of Alzheimer’s dementia[3, 4]. As a result, the heterogeneous and multifactorial nature of this complex disease is increasingly appreciated.

Post-traumatic stress disorder (PTSD) is a psychological disorder that arises from exposure to traumatic events such as combat, assault, or natural disasters[5]. The characteristic clinical manifestations of PTSD cover mode disturbances, behavioral changes, somatic symptoms, and cognitive impairment[5]. Although PTSD and AD are distinct neuropsychological diseases, recent clinical studies and basic research have implicated potential mechanistic links between the two conditions. Epidemiological studies reported that subjects vulnerable to PTSD are also susceptibility to AD in their later life[6]. In addition, neuroimaging data showed that PTSD and AD overlap in structural changes in multiple brain regions including the hippocampus, which are pivotal for emotional processing and cognitive function[7, 8]. Moreover, shared disturbances in key neurotransmitters such as glutamate and serotonin as well as common deregulation of several molecular pathways such as the actin nucleator Formin 2 (Fmn2)-ERK1/2 signaling and the transcription factor nuclear factor-κB (NF-κB) signaling further support the biological basis linking PTSD and AD[9–14]. Of note, a previous genome-wide pleiotropy analysis using genome-wide association studies (GWAS) datasets from patients with AD and PTSD has identified several shared loci between the two conditions, implicating a role of genetic risks in predisposing AD development from PTSD[15].

GWAS analysis has been widely used to identify the association of pathogenic variants with diseases. However, its limitation to inform the consequences of the identified genetic variants has created chasm that constrains the use of GWAS data to understand the pathogenesis of complex diseases[16]. Transcriptome-wide association Studies (TWAS) is a newly developed tool in computational biology[17]. Tissue-specific TWAS gene expression modeling is dictated by expression quantitative loci (eQLT) data to establish associations between single-nucleotide polymorphism (SNPs) and gene expression changes to account for gene expression differences in addition to the variety of expressed sequences from a tissue type-specific perspective. In view of this feature, TWAS functions as a critical complement to GWAS to demonstrate the causal effects of specific sequences towards phenotypic discrepancies, especially those with established genetic risks. Additional to this benefit, TWAS enables tissue specificity and lowers the computational burden of genome analysis by targeting its analysis towards highly relevant gene sequences. As a result of these factors, TWAS is a highly adaptive and effective genome analysis tool, which has been used in investigations of various complex traits and diseases.

In this study, we performed hippocampal TWAS analysis by using GWAS summary statistics from large-scale AD and PTSD cohorts. At the nominal significance of 0.05, 120 PTSD/AD- overlapped susceptibility genes were determined in the hippocampus. Among these PTSD/AD- overlapped susceptibility genes, 68 underwent upregulation and 52 were downregulated. Further functional annotation analysis mounted these PTSD/AD-overlapped susceptibility genes to multiple critical biological pathways, among which metabolism-related pathways were listed on the top. The most parsimonious interpretation of our data is that PTSD and AD share common susceptibility genes. The deregulation of these PTSD/AD-overlapped susceptibility genes may promote disease through disturbances in critical pathways in the hippocampus. Further in-depth experimental investigation will help to delineate the molecular pathways that contribute to both PTSD and AD as well as predispose AD onset from PTSD from a perspective of personalized medicine.

## Results

### Hippocampal TWAS analysis of AD and PTSD cohorts

To explore whether PTSD and AD share common deregulation of gene expression in the hippocampus, we performed hippocampal FUSION transcriptome-wide association studies (TWAS) using GWAS summary statistics from an AD cohort (90,338 cases; 1,036,225 controls) and a PTSD cohort (137,136 cases; 1,085,746 controls)[18–20] (**Fig. 1**). We integrated the expression quantitative trait loci (eQTL) reference panels and linkage disequilibrium (LD) scores. Manhattan plots generated from FUSION TWAS analysis (**supplementary table 1**) summarized susceptibility genes that were associated with PTSD (**Fig. 2A**) and AD (**Fig. 2B**), respectively. Through the analysis using nominal significance at 0.05, we identified 1,422 AD-associated genes (**Fig. 2C**) and 2,145 PTSD-associated genes (**Fig. 2D**).

**Figure 1.**
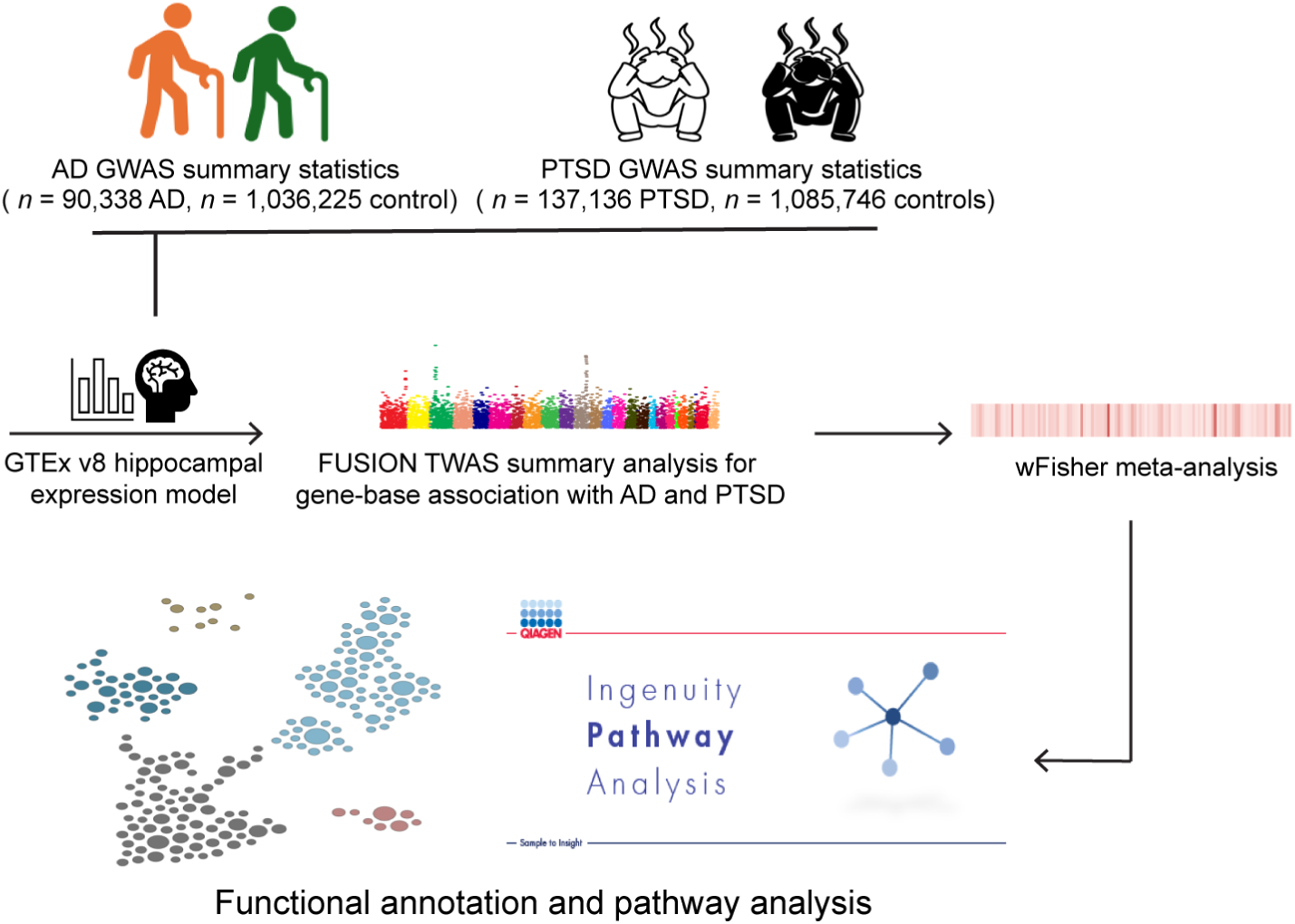
Schematic graph. GWAS: genome-wide association studies, AD: Alzheimer’s disease, PTSD: Post-traumatic stress disorder, wFisher: weight Fisher method.

**Figure 2.**
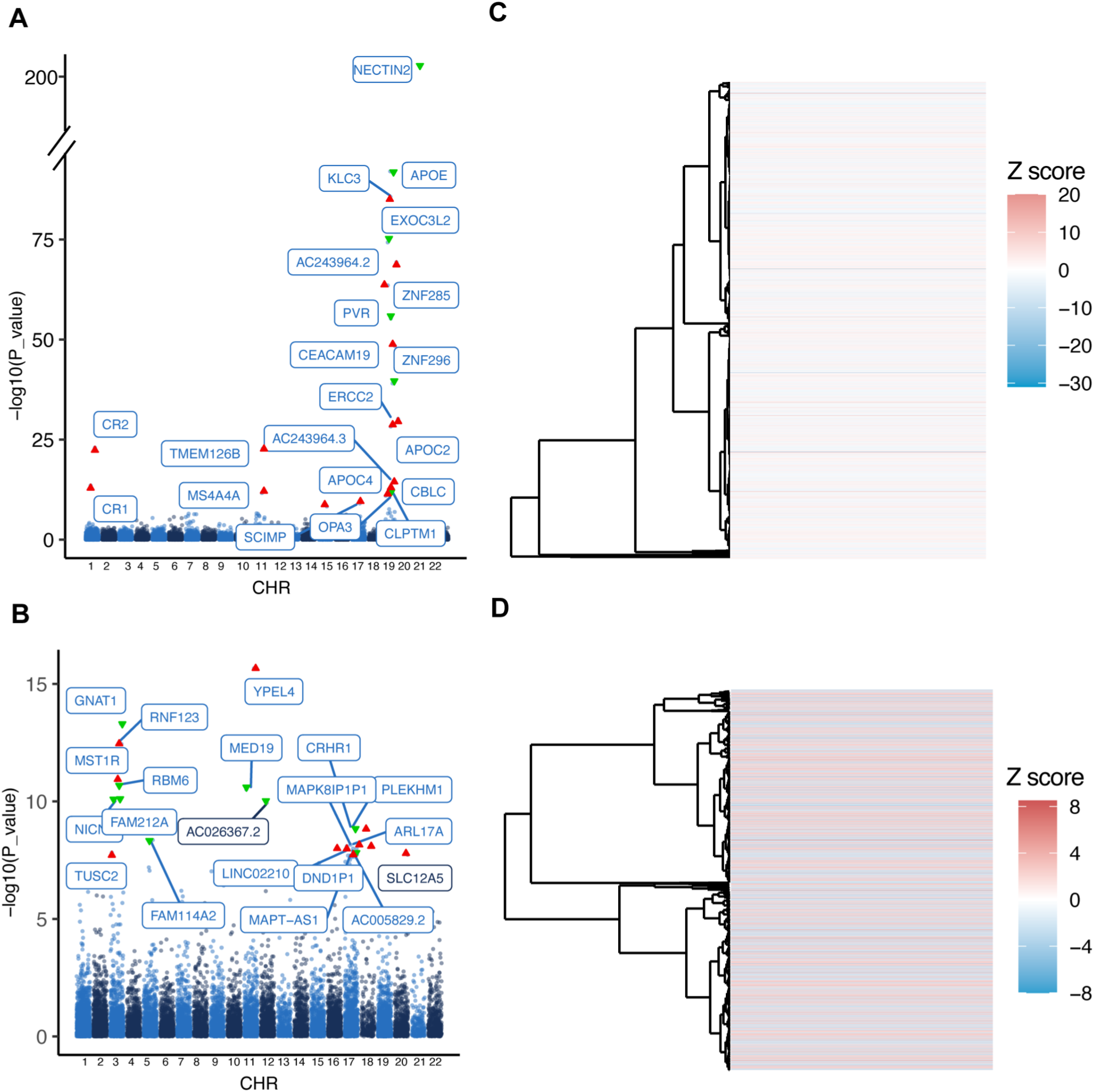
Hippocampal FUSION-TWAS analysis of AD and PTSD GWAS summary data. (**A**) Manhattan plot of AD hippocampal FUSION-TWAS result. *n* = 71,880 AD, 383,378 nonAD controls. (**B**) Clustered heatmap of AD hippocampal TWAS identified genes with *p* < 0.05. (**C**) Manhattan plot of AD hippocampal FUSION-TWAS result. *n* = 137,136 PTSD, 1,085,746 controls. (**D**) Clustered heatmap of PTSD hippocampal TWAS identified genes with *p* < 0.05.

### Cross-disease association analysis of PTSD/AD-overlapped susceptibility genes

To determine the susceptibility genes overlapped in both AD and PTSD, we performed cross-disease association analyses. The FUSION TWAS results were sequentially filtered as shown in the Sankey plot (**Fig. 3A**). At the nominal significance of 0.05, 230 PTSD/AD- overlapped susceptibility genes (**supplementary table 2**) were determined. Afterwards, we performed further filtering process to keep genes with consistent direction of regulation in both diseases. A total of 120 PTSD/AD-overlapped susceptibility genes were determined, among which 68 underwent upregulation and 52 were downregulated (**Fig. 3B-2C**). To enhance statistical power for the combined *p*-values, we performed a meta-analysis using the weighted Fisher’s method (wFisher) based on TWAS summary statistics from the AD and PTSD cohorts (**Fig. 3D**). These findings indicate that PTSD and AD have shared genetic risks that affect the transcriptomic landscape in brain regions vulnerable to both diseases.

**Figure 3.**
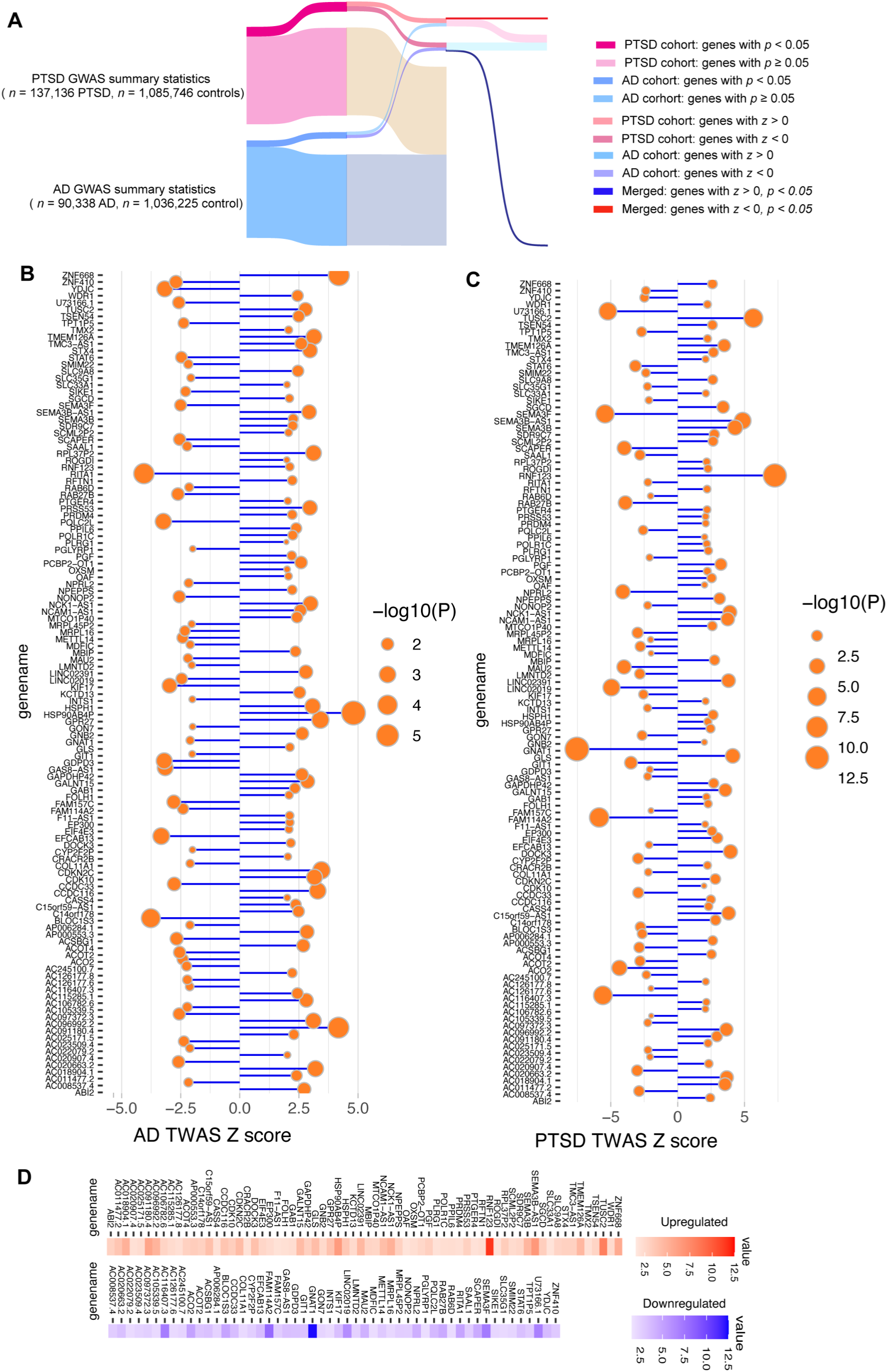
Cross-disease susceptibility genes identification. (**A**) Sankey plot to represent hippocampal AD/PTSD TWAS analysis result filtration for susceptibility genes. (**B&C**) Lollipop graphs for AD (**B**) and PTSD (**C**) overlapped significant gene with same expression direction. *P* < 0.05. (**D**) wFisher meta analysis for AD/PTSD susceptibility genes. Upper panel represents upregulated genes, lower panel represents downregulated genes.

### Functional annotation of cross-disease susceptibility genes

To further investigate potential molecular links between the two diseases, we employed ingenuity pathway analysis (IPA) for comparative analysis of canonical pathways using the AD/PTSD-overlapped susceptibility genes as identified in **Fig. 3B&C**. In the hippocampus, these 120 genes were enriched across several overlapping pathways, especially those related to metabolic functions (**Fig. 4**). We performed canonical pathway analysis based on the wFisher meta-analysis results and that revealed enrichment in several critical metabolic processes such as “Acyl-CoA Hydrolysis”, “Fatty Acid Biosynthesis Initiation”, “Glutamine Degradation”, and “Fatty Acid Activation”. (**Fig. 4A**). For a better understanding of the impacts of AD and PTSD overlapped susceptibility genes on hippocampal functions, we employed the hippocampal functional gene network analysis in HumanBase (https://hb.flatironinstitute.org)[21] and mounted the 120 AD and PTSD shared susceptibility genes to key biological processes in the hippocampus, including “developmental growth” (Module 2) and “protein localization” (Module 3) (**Fig. 4B**). Put together, these results highlight shared alterations in critical pathways related to pivotal biological processes, especially cell metabolism in the hippocampus across AD and PTSD.

**Figure 4.**
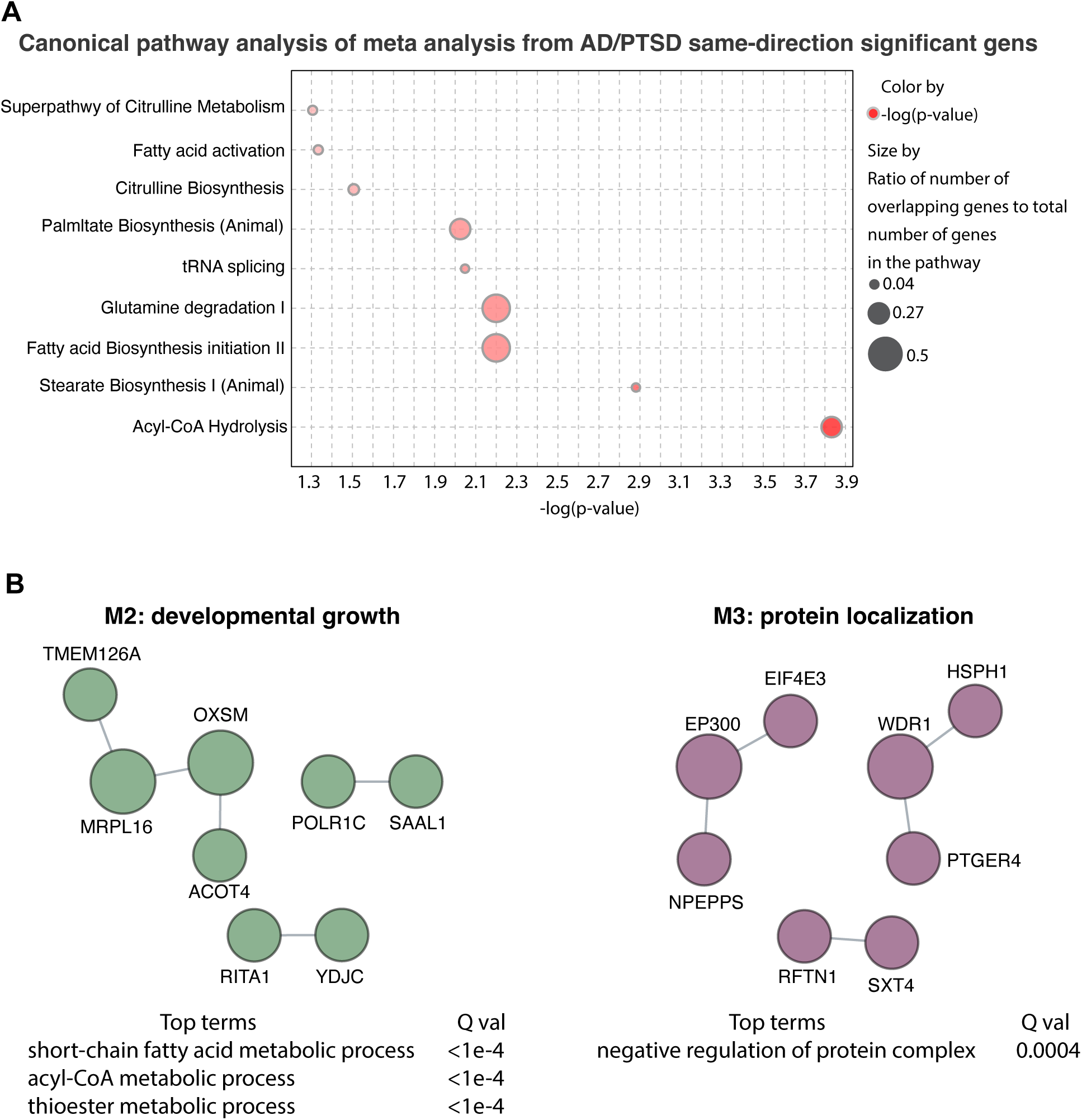
Cross-disease functional analysis. (**A**) Cross-disease IPA canonical pathway identification using AD/PTSD wFisher meta analysis result. (**B**) AD and PTSD shared biological process analysis.

### Expression of AD/PTSD-sensitive metabolic pathway-related genes in the hippocampus

Different cell types express distinct transcriptomic landscape. Therefore, it is of significance to examine cell type-specific gene expression. To further investigate how metabolic pathways influence brain function in AD and PTSD, we examined the expression patterns of key genes involved in the pathways identified in **Fig. 4B** using the CZ CELLxGENE platform (https://cellxgene.cziscience.com/docs/01CellxGene)[22]. The results showed that these genes were broadly expressed across various brain cell types, including neurons, neural stem cells, oligodendrocytes, microglia, astrocytes, and brain vascular cells (**Fig. 5**). In the hippocampus, the genes were also expressed in multiple hippocampal cell types such as hippocampal pyramidal neurons, granule cells, interneurons, and astrocytes except for Acyl-CoA Thioesterase 4 (ACOT4), which was not expressed in hippocampal pyramidal neurons. The widespread expression of these AD/PTSD-sensitive genes highlights the broad impacts of altered expression of these genes on hippocampal function.

**Figure 5.**
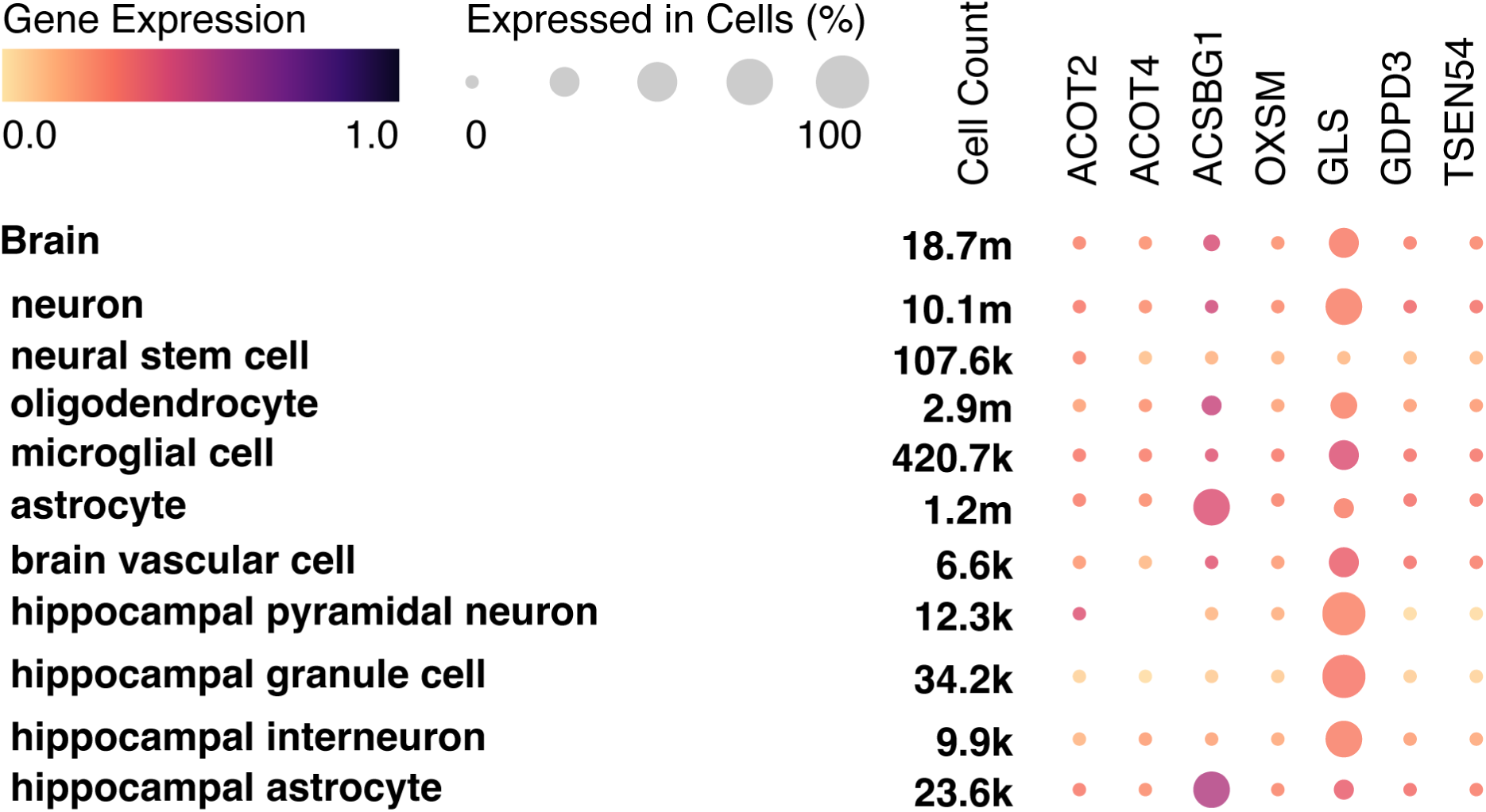
Cell-type-specific expression profiling of shared AD/PTSD metabolic pathway– related genes in the brain. Expression pattern of IPA-identified AD/PTSD overlapped metabolic pathway-related genes in multiple brain cell types and different hippocampal cells.

## Discussion

An association between post-traumatic stress disorder (PTSD) and Alzheimer’s disease (AD) has been repeatedly implicated in previous clinical and basic research. However, the precise mechanisms linking the two neuropsychiatric conditions have not yet been elucidated. A previous study using GWAS dataset has identified multiple loci and pathogenic variants overlapping in both PTSD and AD[15], which provides critical evidence of the genetic link between the two diseases. Tissue-specific TWAS is a recently developed method that has its capacity to complement GWAS analysis in a better understanding of the impact of pathogenic variants on gene regulation in a tissue-specific manner. Inspired by the advantages of tissue-specific TWAS, in this study, we performed TWAS analysis using GWAS summary statistics from large-scale cohorts of PTSD and AD and examined the shared genetic risks between PTSD and AD and their impacts on hippocampus gene expression in these two conditions. In addition to the identified PTSD/AD-overlapped susceptibility genes, our further functional annotation mounted these genetic risks to multiple biological pathways, especially those related to key metabolic regulations. It is consensually accepted that metabolic homeostasis is important to synaptic strength and brain function[23, 24]. As a result, disruptions in metabolism have been consistently observed patients with neurological disorders including PTSD and AD[25–28]. AD patients frequently report metabolic changes before cognitive decline[29]. Previous studies have firmly established the pivotal role of metabolic perturbations including insulin resistance, mitochondrial dysfunction and cholesterol and sphingolipid dysmetabolism in the etiopathogenesis of AD, which promotes the appraisal of metabolic pathways of this neurodegenerative disorder[30–34]. Of note, metabolic conditions such as obesity, dyslipidemia, type 2 diabetes, and mitochondrial dysfunction are also well-documented risks associated with PTSD[27, 35–37]. A previous study showed a close association between metabolic syndrome and cortical atrophy in military veterans with PTSD[38], implicating a contribution of dysmetabolism to PTSD-associated brain structural changes. Despite the agreement that systematic and brain dysmetabolism are involved in the pathogenesis of both AD and PTSD, there exists a debate over the causal role of dysmetabolism in the development of these disorders. Previous studies frequently define dysmetabolism in PTSD and AD as secondary responses to other disease-promoting factors such as chronic stress, inflammatory reactions, and the hypothalamic-pituitary-adrenal (HPA) axis instability[39–42]. Although we cannot refute the possibility that dysmetabolism is an aftereffect or adaptive changes in patients with PTSD and AD, our findings of the deleterious impact of PTSD/AD-overlapped susceptibility gene regulation on key metabolic pathways, at least in the hippocampus, implicate an alternative mechanism. In view of the detrimental effect of dysmetabolism on brain function and the pivotal role of the hippocampus in cognition and emotion[43–45], we thus cautiously formulate a hypothesis that subjects carrying genetic risks may gradually develop hippocampal dysmetabolism, which compromises neuronal function, rendering susceptibility to failed defense strategies in the context of stress, culminating in increased risk of PTSD as well as AD in their later life. To this end, at least a subgroup of PTSD and AD patients, who carry the determined susceptibility genes, converge in hippocampal dysmetabolism in their disease development as well as the vicious sequence of PTSD to AD. Further in-depth investigations using experimental approaches are in immediate need to address this critical scientific question.

Another critical finding merits discussion is that our TWAS analysis determined alterations in the expression of ACOT2 and ACOT4 in the hippocampus of both AD and PTSD, resulting in acyl-CoA hydrolysis as the top pathway affected in both AD and PTSD. Acyl-CoA hydrolysis involves the breakdown of acyl-CoA into free fatty acids and coenzyme A, a reaction catalyzed by acyl-CoA thioesterases (ACOTs) [46, 47]. Our findings of the association of hippocampal ACOT2 and ACOT4 deregulation with the risk of both AD and PTSD implicates a contribution of fatty acid dysmetabolism to the development of the two disorders, corroborating previous reports of lipid dysmetabolism in the pathogenesis of both AD and PTSD [48–50]. Noteworthy, both ACOT2 and ACOT4, the two key enzymes in fatty acid metabolism, are expressed in various types of brain cells. Previous studies indicate that fatty acid oxidation is pivotal for the functions of glial cells including astrocytes, microglia, and oligodendrocytes. Consistent with the role of astrocytes as the frontline of fatty acid transport from the peripheral circulation into the brain, astrocytes are active in using fatty acids to produce energy, synthesize proteins as well as facilitate neurotransmitter production via astrocyte-neuron interactions [51, 52]. Disrupted lipid clearance, particularly in the astrocytes, results in lipid accumulation, contributing to the formation of amyloid-beta (Aβ) plaques and tau tangles, the defining pathological features of AD [28, 53]. Moreover, microglial cells use fatty acids as an alternative energy source to meet the high energy demand during microglial activation, and deregulated lipid usage may underlie microglial abnormalities [54]. In oligodendrocytes, which generate the lipid-rich myelin sheath, impaired lipid metabolism can lead to white matter abnormalities, a common feature observed in both AD and PTSD [55–57]. Furthermore, abnormal lipid accumulation can compromise brain vasculature, induce insulin resistance, and disrupt neuronal membrane integrity, further contributing to neurodegeneration[58, 59]. However, despite the importance of fatty acid oxidation through acyl-CoA hydrolysis to glial and vascular cell functions, the current opinion is that neurons have limited capability in using fatty acids to fuel neuronal activity[60], which brings up a scientific question of whether and how altered acyl-CoA hydrolysis affects neuronal function. Of note, neural stem/progenitor cells give rise to granule neurons[60]. Previous studies indicate that mitochondrial fatty acid β-oxidation is important to the development of neural progenitor cells, and the defects of mitochondrial fatty acid β-oxidation may underlie developmental brain disorders such as Autism [61]. To this end, if we could perform a thought experiment, there exists a possibility that downregulation of ACOT2 in neural progenitor cells may impair the development and maturation of hippocampal pyramidal neurons, which dampens the reserve of neuronal activity, culminating in augmented sensitivity of affected subjects to the development of PTSD and AD in sequence. This hypothesis is further supported by our findings from the hippocampal functional gene network analysis, which enriched in “developmental growth”. Lastly, our TWAS analysis showed a negative relationship between ACOT2 expression and AD/PTSD risk. In contrast, AD and PTSD are positively associated with ACOT4 expression. ACOT2 primarily resides in mitochondria and is involved in mitochondrial β-oxidation of branched chain fatty acid, while ACOT4 is abundant in peroxisomes and contributes to peroxisomal degradation of very long chain fatty acids [47]. In this regard, the opposing effects of ACOT2 and ACOT4 deregulation on brain dysfunction in PTSD and AD remain unresolved. This discrepancy may arise from the versatile metabolic products of peroxisomal very long chain fatty acids [47]. On a related note, elevated very long chain fatty acids and dysregulated peroxisomal very long chain fatty acid metabolism has been previously linked to neurotransmission defects and pathological characteristics of AD including brain amyloidosis and tauopathy [62, 63], further supporting impaired homeostasis of very long chain fatty acid metabolism in potentiating neuronal damages. These outstanding questions warrant further experimental investigations.

The limitation of this study should also be noted, that is, the underrepresentation of racially and ethnically diverse populations. The large-scale GWAS summary statistics for AD and PTSD used in our TWAS analysis were primarily derived from individuals of European ancestry, which may constrain the generalizability of our findings to other populations. This lack of diversity can mask population-specific genetic risk factors, environmental exposures, and disease trajectories. Future studies will incorporate cohorts from a broader range of ancestral backgrounds to better elucidate the genetic links between AD and PTSD across diverse populations. Another limitation of this proof-of-concept study is its reliance solely on computational analysis to identify genetic links between AD and PTSD, without experimental validation. As a next step, we plan to use these findings to inform experimental designs that will test and validate computational predictions. In addition, longitudinal studies using PTSD cohorts to assess the role of these identified susceptibility genes and pathogenic variants in promoting AD development will be informative to assess the potential of the computational findings in early diagnosis and prevention of PTSD, AD, as well as AD conversion from PTSD patients.

In summary, we identified shared gene variants in brain regions vulnerable to both AD and PTSD, specifically the hippocampus. These genes were enriched in key metabolic pathways, especially in the key pathway of fatty acid metabolism, which align with existing evidence from both patient studies and animal models. Although in *vivo* experimental studies are needed to further explore the detailed impact of these genes on neuronal and brain function, our results reveal gene variant–regulated metabolic pathways as a novel molecular link between AD and PTSD. These pathways not only shed light on shared disease mechanisms but also hold promise as biomarkers and potential personalized therapeutic targets, particularly for PTSD-associated risk of developing AD.

## Materials and Methods

### Hippocampal FUSION TWAS analysis

To explore the relationship between single nucleotide polymorphism (SNP)-associated phenotypes and gene expression in the hippocampus, we performed a transcriptome-wide association study (TWAS) using the FUSION-TWAS method [18]. We obtained freely accessible genome-wide association study (GWAS) summary data for both Alzheimer’s disease cohort including 71,880 AD cases and 383,378 non-AD controls and Post-traumatic stress disorder (PTSD) including 137,136 cases and 1,085,746 controls from the original publication [19, 20]. Cis-expression quantitative trait loci (cis-eQTL) weights for the hippocampus and reference linkage disequilibrium (LD) covariance data for chromosomes 1–22 calculated based on GTEx V8 dataset were obtained via the FUSION-TWAS github [18]. Hippocampal Gene expression imputation was carried out in accordance with the FUSION association test protocol.

### Functional annotation of the identified genes

QIAGEN Ingenuity Pathway Analysis (IPA) was used to perform comparative analysis and identify canonical pathways. To identify shared susceptibility genes, we filtered the AD/PTSD TWAS results for genes with *P* < 0.05 in both disorders and used this list for IPA analysis. We first conducted a comparison analysis to uncover shared disease-related pathways and functional associations using genes exhibiting the same direction of expression changes (based on FUSION TWAS z-scores) and significant p value (*P* < 0.05) in both AD and PTSD. We then applied wFisher meta-analysis p-values to further explore canonical pathways implicated in both AD and PTSD. Lastly, to further investigate the biological processes regulated by these overlapping genes, we carried out hippocampal gene network analysis using HumanBase (https://hb.flatironinstitute.org), focusing on genes with consistent expression direction between AD and PTSD patients[21].

### Tissue and cell gene expression screening

To investigate the expression patterns of genes that are involved in IPA determined AD/PTSD shared metabolic pathways, we leveraged open-access human single cell gene expression data available through the Cell Types Database on CZ CELL x GENE [22]. Genes that are essential to AD/PTSD-related metabolic pathways were obtained from IPA analysis result. The screening included multiple brain cell types, such as neurons, neural stem cells, oligodendrocytes, microglia, astrocytes, brain vascular cells, as well as hippocampal pyramidal neurons, granule cells, interneurons, and astrocytes. Both gene expression levels and the percentage of expressing cells were assessed in this analysis.

### Statistical analysis and meta-analysis

All data were analyzed using Prism graphpad 9 statistical software unless otherwise indicated. The meta-analysis of TWAS statistics from the two cohorts was performed with the use of the weighted Fisher’s method (wFisher) in metapro R package [64]. *P*-values from the TWAS analysis, along with corresponding sample sizes and effect directions, were used as input for the meta-analysis to calculate combined p-values. Genes with a combined *p*-value less than 0.05 and consistent *z*-score direction across both cohorts were considered transcriptome-wide significant. Graphs are generated using R packages and HumanBase, CZ CELL x GENE web services.

## Data Availability

All original data are available upon request from the corresponding author.

